# Identifying the measurements required to estimate rates of COVID-19 transmission, infection, and detection, using variational data assimilation

**DOI:** 10.1101/2020.05.27.20112987

**Authors:** Eve Armstrong, Manuela Runge, Jaline Gerardin

## Abstract

We demonstrate the ability of statistical data assimilation to identify the measurements required for accurate state and parameter estimation in an epidemiological model for the novel coronavirus disease COVID-19. Our context is an effort to inform policy regarding social behavior, to mitigate strain on hospital capacity. The model unknowns are taken to be: the time-varying transmission rate, the fraction of exposed cases that require hospitalization, and the time-varying detection probabilities of new asymptomatic and symptomatic cases. In simulations, we obtain accurate estimates of undetected (that is, unmeasured) infectious populations, by measuring the detected cases together with the recovered and dead - and without assumed knowledge of the detection rates. These state estimates require a measurement of the recovered population, and are tolerant to low errors in that measurement. Further, excellent estimates of all quantities are obtained using a temporal baseline of 112 days, with the exception of the time-varying transmission rate at times prior to the implementation of social distancing. The estimation of this transmission rate is sensitive to contamination in the data, highlighting the need for accurate and uniform methods of reporting. Finally, we employ the procedure using real data from Italy reported by Johns Hopkins. The aim of this paper is not to assign extreme significance to the results of these specific experiments *per se*. Rather, we intend to exemplify the power of SDA to determine what properties of measurements will yield estimates of unknown model parameters to a desired precision - all set within the complex context of the COVID-19 pandemic.

## I. INTRODUCTION

The coronavirus disease 2019 (COVID-19) is burdening health care systems worldwide, threatening physical and psychological health, and desecrating the global economy. It is our nation’s top priority to assuage this harm. In particular, it is vital to maintain appropriate social behavior to avoid straining hospital capacity, and meanwhile minimize the psychological stress that social restrictions place upon citizens. Given the uncertainties, however, in the intrinsic properties of the disease and inaccuracies in the recording of its effects on the population [1–3], an optimal protocol for social behavior is far from clear. It is invaluable to identify any and all methodologies that may be brought to bear upon this situation.

Within this context, we seek a means to quantify what data must be recorded in order to estimate specific unknown quantities in an epidemiological model tailored to the COVID-19 pandemic. These unknown quantities are: i) the transmission rate, ii) the fraction of the exposed population that acquires symptoms sufficiently severe to require hospitalization, and iii) time-varying detection probabilities of asymptomatic and symptomatic cases. In this paper, we demonstrate the ability of statistical data assimilation (SDA) to quantify the accuracy to which these parameters can be estimated, given certain properties of the data including noise level.

SDA is an inverse formulation [4]: a machine learning approach designed to optimally combine a model with data. Invented for numerical weather prediction [5–10], and more recently applied to biological neuron models [11–17], SDA offers a systematic means to identify the measurements required to estimate unknown model parameters to a desired precision.

Data assimilation has been presented as a means for general epidemiological forecasting [18], and one work has examined variational data assimilation specifically - the method we employ in this paper - for estimating parameters in epidemiological models [19]. Related Bayesian frameworks for estimating unknown properties of epidemiological models have also been explored [20,21]. To date, there have been two employments of SDA for COVID-19 specifically. Ref [22] used a simple SIR (susceptible/infected/recovered) model, and Ref [23] expanded the SIR model to include a compartment of patients in treatment.

Two features of our work distinguish this paper as novel. First, we significantly expand the model in terms of the number of compartments. The aim here is to capture key features of COVID-19 so as to eventually inform state policy on containing the pandemic. These features are: i) asymptomatic versus symptomatic populations, ii) undetected versus detected cases, and iii) two hospitalized populations: those who do and do not require critical care. For our motivations for these choices, see *Model*. Second, we employ SDA for the specific purpose of examining the sensitivity of estimates of time-varying parameters to various properties of the measurements, including the degree of noise (or error) added. Moreover, we aim to demonstrate the power and versatility of the SDA technique to explore what is required of measurements to complete a model with a dimension sufficiently high to capture the complexities of COVID-19 - an examination that has not previously been done.

To this end, we sought to estimate the parameters noted above, using simulated measurements representing a metropolitan-area population loosely based on New York City. We examined the sensitivity of estimations to: i) the subpopulations that were sampled, ii) the temporal baseline of sampling, and iii) uncertainty in the sampling.

Our findings using simulated data are threefold. First, reasonable estimations of time-varying detection probabilities require the reporting of new detected cases (asymptomatic and symptomatic), dead, and recovered, and low (~ ten percent) noise is well tolerated. Second, the information contained in the measured *detected* populations propagates successfully to the estimation of the numbers of *undetected* cases entering hospitals. Third, a temporal baseline of 112 days (the recent past) is sufficiently long for the SDA procedure to capture the general trends in the evolution of the model populations, the detection probabilities, and the time-varying transmission rate following the implementation of social distancing. The transmission rate at early times is highly sensitive to contamination in the measurements and to the temporal baseline of sampling. Finally, we test the procedure on real data, using the reports from Italy that are listed in the open-access github repository of the Johns Hopkins project Systems Science and Engineering [24].

The key sections to note in this paper are: *The experiments* - which describes the studies done and our motivations for them, *Results: General Findings*, and *Conclusion*.

Finally, we comment on the model parameters and initial conditions that are taken in this paper to be known quantities. As noted, our aim is to exemplify SDA as a tool to examine how certain properties of data impact the estimation of unknown model parameters. As any tool of possible utility must be brought to bear upon this situation in a timely manner, we made some assumptions that do not reflect all that is known to date about the COVID-19 pandemic. It is important to test the procedure for robustness over various parameter ranges, “correct” or otherwise. Meanwhile, we are testing the procedure’s stablity over a wide range of choices for parameter values and initial conditions, including those that reflect most accurately the current state of our knowledge of the pandemic. Moreover, is our hope that the exercises described in this paper can be taken and applied to a host of complicated questions surrounding COVID-19.

## II. MODEL

The model is written in 22 state variables, each representing a subpopulation of people; the total population is conserved. Figure 1 shows a schematic. Each member of a Population *S* that enters an Exposed Population (*E*) ultimately reaches either Recovered (*R*) or Dead (*D*). Absent additive noise, the model is deterministic. The red ovals indicate the variables that will correspond to measured quantities in the inference experiments of this paper.

**Figure 1:**
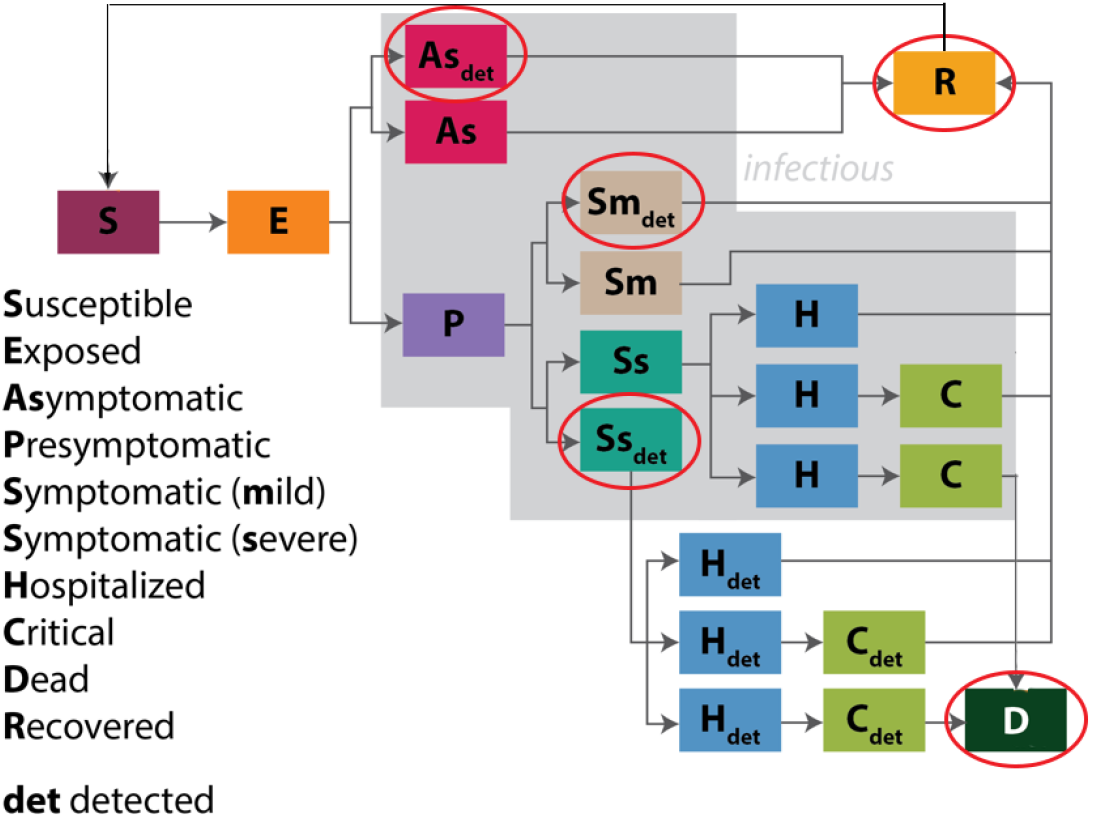
Schematic of the model. Each rectangle represents a population. Note the distinction of asymptomatic cases, undetected cases, and the two tiers of hospitalized care: *H* and *C*. The aim of including this degree of resolution is to inform policy on social behavior so as to minimize strain on hospital capacity. The red ovals indicate the variables that correspond to measured quantities in the inference experiments of this paper.

As noted, the model is written with the aim to inform policy^1^ on social behavior so as to avoid overwhelming hospital capacity. To this end, the model resolves asymptomatic-versus-symptomatic cases, undetected-versus-detected cases, and the two tiers of hospitalization: the general *H* versus the critical care *C* populations.

The resolution of asymptomatic versus symptomatic cases was motivated by an interest in what requirements exist to control the epidemic. For example, is it sufficient to focus only on symptomatic individuals, or must we also target and address asymptomatic individuals who are not personally suffering from disease?

The detected and undetected populations exist for two reasons. First, we seek to account for underreporting of cases and deaths. Second, we will eventually seek a model structure that can simulate the impact of increasing detection rates on disease transmission, including the impact of contact tracing. Thus the model was structured from the beginning so that we might examine the effects of interventions that were imposed later on. The ultimate aim here is to inform policy on the requirements for containing the epidemic.

We included both *H* and *C* populations because hospital inpatient and ICU bed capacities are the key health system metrics that we aim to avoid straining. Any policy that we consider must include predictions on inpatient and ICU bed needs. Preparing for those needs is a key response need if or when the epidemic grows uncontrolled.

Note on Figure 1 the return arrow from the recovered *R* to susceptible *S* population. The return of any portion of *R* to *S* is believed to be zero. Nevertheless, we assayed the ability of SDA to assimilate some portion of *R* back into *S*, as a proof-of-principle that it can be done readily. For details of the model, including the reaction equations and descriptions of all state variables and parameters, see *Appendix A*.

## III. METHOD

### A. General inference formulation

SDA is an inference procedure, or a type of machine learning, in which a model dynamical system is assumed to underlie any measured quantities. This model ***F*** can be written as a set of *D* ordinary differential equations that evolve in some parameterization *t* as:

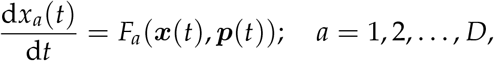

where the components *x_a_* of the vector x are the model state variables, and unknown parameters to be estimated are contained in ***p***(*t*). A subset *L* of the *D* state variables is associated with measured quantities. One seeks to estimate the *p* unknown parameters and the evolution of all state variables that is consistent with the *L* measurements.

A prerequisite for estimation using real data is the design of simulated experiments, wherein the true values of parameters are known. In addition to providing a consistency check, simulated experiments offer the opportunity to ascertain *which* and *how few* experimental measurements, in principle, are necessary and sufficient to complete a model.

### B. Optimization framework

SDA can be formulated as an optimization, wherein a cost function is extremized. We take this approach, and write the cost function in two terms: 1) one term representing the difference between state estimate and measurement (measurement error), and 2) a term representing model error. It will be shown below in this Section that treating the model error as finite offers a means to identify whether a solution has been found within a particular region of parameter space. This is a non-trivial problem, as any nonlinear model will render the cost function non-convex. We search the surface of the cost function via the variational method, and we employ a method of annealing to identify a lowest minumum - a procedure that has been referred to loosely in the literature as variational annealing (VA).

The cost function *A*_0_ used in this paper is written as:

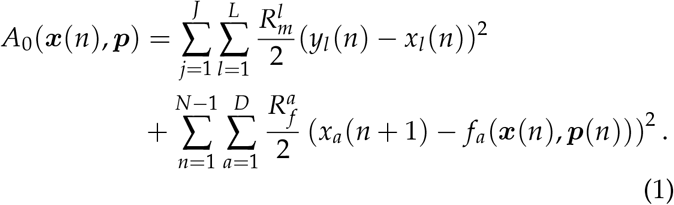

One seeks the path ***X***^0^ = *x*(0),…,*x*(*N*),***p***(*0*),…***p***(*N*) in state space on which *A*_0_ attains a minimum value^2^. Note that Equation 1 is shorthand; for the full form, see *Appendix A* of Ref [17]. For a derivation - beginning with the physical Action of a particle in state space - see Ref [26].

The first squared term of Equation 1 governs the transfer of information from measurements *y_l_* to model states *x_l_*. The summation on *j* runs over all discretized timepoints *J* at which measurements are made, which may be a subset of all integrated model timepoints. The summation on *l* is taken over all *L* measured quantities.

The second squared term of Equation 1 incorporates the model evolution of all *D* state variables *x_a_*. The term *f_a_*(*x*(*n*)) is defined, for discretization, as: 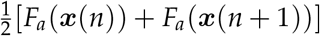. The outer sum on *n* is taken over all discretized timepoints of the model equations of motion. The sum on *a* is taken over all *D* state variables.

*R_m_* and *R_f_* are inverse covariance matrices for the measurement and model errors, respectively. We take each matrix to be diagonal and treat them as relative weighting terms, whose utility will be described below in this Section.

The procedure searches a (*D* (*N* + 1) + *p* (*N* + 1))-dimensional state space, where *D* is the number of state variables, *N* is the number of discretized steps, and *p* is the number of unknown parameters. To perform simulated experiments, the equations of motion are integrated forward to yield simulated data, and the VA procedure is challenged to infer the parameters and the evolution of all state variables - measured and unmeasured - that generated the simulated data.

This specific formulation has been tested with chaotic models [27–30], and used to estimate parameters in models of biological neurons [12,13,15,17,31,32], as well as astrophysical scenarios [33].

### C. Annealing to identify a solution on a non-convex cost function surface

Our model is nonlinear, and thus the cost function surface is non-convex. For this reason, we iterate - or anneal - in terms of the ratio of model and measurement error, with the aim to gradually freeze out a lowest minimum.

This procedure was introduced in Ref [34], and has since been used in combination with variational optimization on nonlinear models in Refs [10, 17, 31, 33] above. The annealing works as follows.

We first define the coefficient of measurement error *R_m_* to be 1.0, and write the coefficient of model error *R_f_* as: 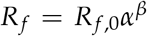, where *R_f_*_,0_ is a small number near zero, *α* is a small number greater than 1.0, and *β* is initialized at zero. Parameter *β* is our annealing parameter. For the case in which *β =* 0, relatively free from model constraints the cost function surface is smooth and there exists one minimum of the variational problem that is consistent with the measurements. We obtain an estimate of that minimum. Then we increase the weight of the model term slightly, via an integer increment in *β*, and recalculate the cost. We do this recursively, toward the deterministic limit of *R_f_* ≫ *R_m_*. The aim is to remain sufficiently near to the lowest minimum to not become trapped in a local minimum as the surface becomes resolved. We will show in *Results* that a plot of the cost as a function of *β* reveals whether a solution has been found that is consistent with both measurements and model.

## IV. THE EXPERIMENTS

### A. Simulated experiments

We based our simulated locality loosely on New York City^3^, with a population of 9 million^4^. Simulations ran from an initial time *t*_0_ of four days prior^5^ to 2020 March 1, the date of the first reported COVID-19 case in New York City [35]. At time *t*_0_, there existed one detected symptomatic case within the population of 9 million^6^.

We chose five quantities as unknown parameters to be estimated (Table 1): 1) the time-varying transmission rate *K_i_*(*t*); 2) the detection probability of mild symptomatic cases *d_Sym_*(*t*), 3) the detection probability of severe symptomatic cases *d_Sys_*(*t*), 4) the fraction of cases that become symptomatic *f_sympt_*, and 5) the fraction of symptomatic cases that become severe enough to require hospitalization *f_severe_*. Here we summarize the key features that we sought to capture in modeling these parameters; for their mathematical formulatons, see *Appendix B*.

**Table 1:** Unknown parameters to be estimated. *K_i_*, *d_Sym_*, and *d_Sys_* are taken to be time-varying. Parameters *f_sympt_* and *f_severe_* are constant numbers, as they are assumed to reflect an intrinsic property of the disease. The detection probability of asymptomatic cases is taken to be known and zero.

| Parameter | Description |
| --- | --- |
| $K_i(t)$ | Time-varying transmission rate |
| $d_{sym}(t)$ | Time-varying detection probability of mild symptomatics |
| $d_{sys}(t)$ | Time-varying detection probability of symptomatics requiring hospitalization |
| $f_{sympt}$ | Fraction of positive cases that produce symptoms |
| $f_{severe}$ | Fraction of symptomatics that are severe |

The transmission rate *K_i_* (often referred to as the effective contact rate) in a given population for a given infectious disease is measured in effective contacts per unit time. This may be expressed as the total contact rate multiplied by the risk of infection, given contact between an infectious and a susceptible individual. The contact rate, in turn, can be impacted by amendments to social behavior^7^.

As a first step in applying SDA to a high-dimensional epidemiological model, we chose to condense the significance of *K_i_* into a relatively simple mathematical form. We assumed that *K_i_* was constant prior to the implementation of a social-distancing mandate, which then effected a rapid transition of *K_i_* to a lower constant value. Specifically, we modeled *K_i_* as a smooth approximation to a Heaviside function that begins its decline on March 22, the date that the stay-at-home order took effect in New York City [39]: 25 days after time *t*_0_. For further simplicity, we took *K_i_* to reflect a single implementation of a social distancing protocol, and adherence to that protocol throughout the remaining temporal baseline of estimation.^8^

Detection rates impact the sizes of the subpopulations entering hospitals, and their values are highly uncertain [2, 3]. Thus we took these quantities to be unknown, and - as detection methods will evolve - time-varying. We also optimistically assumed that the methods will improve, and thus we described them as increasing functions of time. We used smoothly-varying forms, the first linear and the second quadratic, to preclude symmetries in the model equations. Meanwhile, we took the detection probability for asymptomatic cases (*d_As_*) to be known and zero, a reasonable reflection of the current state of testing.

Finally, we assigned as unknowns the fraction of cases that become symptomatic (*f_sympt_*) and fraction of symptomatic cases that become sufficiently severe to require hospitalization (*f_severe_*), as these fractions possess high uncertainties (Refs [40] and [41], respectively). As they reflect an intrinsic property of the disease, we took them to be constants. All other model parameters were taken to be known and constant^9^ (*Appendix A*).

The simulated experiments are summarized in the schematic of Figure 2. They were designed to probe the effects upon estimations of three considerations: a) the number of measured subpopulations, b) the temporal baseline of measurements, and c) contamination of measurements by noise. To this end, we designed a “base” experiment sufficient to yield an excellent solution, and then four variations on this experiment.

**Figure 2:**
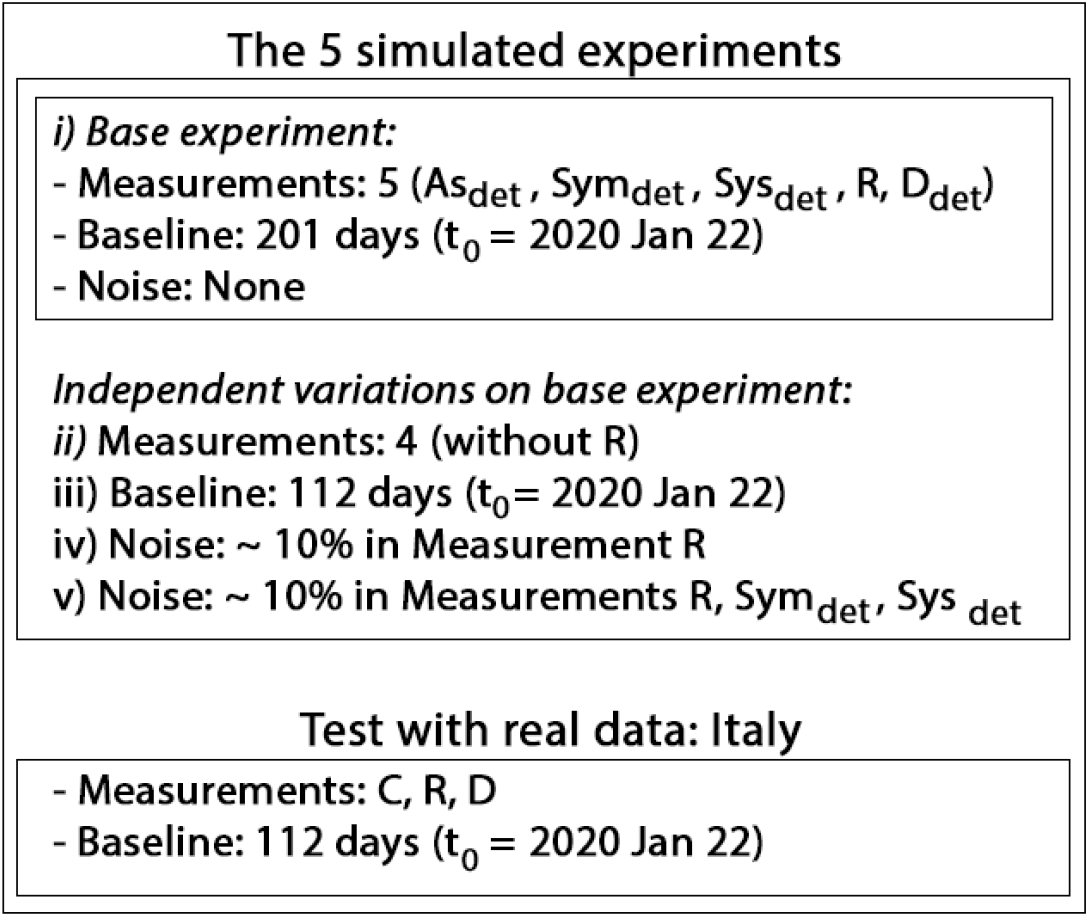
Schematic of the inference experiments. For Italy, *C* is distributed among the model variables *As_det_, Sym_det_*, *Sys_det_*, for correspondence with the model.

The base experiment (*i* in Figure 2) possesses the following features: a) five measured populations: detected asymptomatic *As_det_*, detected mild symptomatic *Sym_det_*, detected severe symptomatic *Sys_det_*, Recovered *R*, and Dead *D*; b) a temporal baseline of 201 days, beginning on 2020 February 26; c) no noise in measurements.

The four variations on this basic experiment (*ii* through *ν* in Figure 2), incorporate the following independent changes. In Experiment *ii*, the *R* population is not measured - an example designed to reflect the current situation in some localities (e.g. Refs [2,3]).

Experiment *iii* reduces the temporal baseline from 201 to 112 days, ending on 2020 May 12: the recent past. The motivation for this design is to ascertain whether - in principle - it was possible to attain an accurate solution that early, given perfectly-recorded data.

Experiment *iv* includes a ~ ten percent noise level in the simulated *R* data, and Experiment *v* includes the same noise level in *R*, *Sym_det_*, and *Sys_det_* (for the form of additive noise, see *Appendix C*).

For each experiment, ten independent calculations were initiated in parallel searches, each with a randomly-generated set of initial conditions on state variable and parameter values.

### B. An example using real data: Italy

We tested the VA procedure using real data obtained from the Johns Hopkins repository [24], using as measurements their reported Confirmed cases *C*, Recovered *R*, and Dead *D*, over a 112-day period. Here, *C* is reported as a cumulative measurement, including *R* and *D*.

To formulate a correspondence with our model, we subtracted from *C* the *R* and *D* numbers and divided the remainder among *As_det_*, *Sym_det_*, and *Sys_det_*. We assumed that the data accord with our model, and assigned zero percent of *C* to the *As_det_* population. Then, absent any well-informed guideline, we assigned 60 and 40% of *C* to *Sym_det_* and *Sys_det_*, respectively. Also in accordance with our model, we multiplied the reported value of *R* by 0.9, as the model assumes a 10% rate of re-entry from the *R* to *S* populations. Finally, for the real data we initialized 50 independent calculations in parallel (rather than 10 for the simulated experiments).

For technical details of all experimental designs and implementation, see *Appendix C*.

## V. RESULT

### A. General findings

The salient results for the simulated experiments *i* through *iv* are as follows:

a. (base experiment): Excellent estimate of all - measured and unmeasured - state variables, and all parameters except for *K_i_*(*t*) at times prior to the onset of social distancing;
b. (absent a measurement of Population *R*): Poor estimate of unmeasured states and time-varying parameters; excellent estimate of static parameters;
c. (shorter baseline of 112 days): Excellent estimate of state variables; excellent estimates of static parameters and time-varying detection probabilities; poor estimate of *K_i_*(*t*) over the full baseline;
d. (~ 10% additive noise in *R*): Estimates of state variables, detection probabilities, and static parameters suffer minimally; poor estimate of *K_i_*(*t*);
e. (~ 10% additive noise in R, *Sym_det_*, *Sys_det_*): Estimates of state variables, detection probabilities, and static parameters suffer mildly; poor estimate of *K_i_*(*t*).

Figures of the estimated time evolution of state variables and time-varying parameters are shown in their respective subsections, and the estimates of the static parameters are listed in Table 2.

**Table 2:** Estimates of static parameters *f_sympt_* and *f_severe_*, over the five simulated experiments and the example using data from Italy. With two exceptions, the reported numbers are taken from the annealing iteration with a value of parameter *β* of 37: once the deterministic limit has been reached (see text). The first exception is Experiment *ii*, for which we chose the result corresponding to *β =* 8: before the solution grows unstable exponentially (see Figure 5). The second exception is the case for real data, where we report results at two values of *β* to show that the estimates vary widely over the annealing process. The result *Italy_I_* corresponds to *β =* 20, prior to the strong imposition of model constraints; *Italy_II_* corresponds to *β =* 37, once the deterministic limit is reached. See specific subsections for details of each experiment.

| Experiment | $f_{symp}$ (true: 0.5) | | $f_{severe}$ (true: 0.3) | |
| --- | --- | --- | --- | --- |
|  | Mean | Variance | Mean | Variance |
| *i* | 0.5 | $5 \times 10^{-9}$ | 0.3 | $2 \times 10^{-9}$ |
| *ii* | 0.41 | 0.005 | 0.13 | 0.003 |
| *iii* | 0.5 | $4 \times 10^{-8}$ | 0.3 | $4 \times 10^{-8}$ |
| *iv* | 0.39 | 0.0 | 0.37 | 0.0 |
| *v* | 0.41 | 0.0 | 0.36 | 0.0 |
| *Italy<sub>I</sub>* | 0.39 | 0.0 | 0.62 | 0.0 |
| *Italy<sub>II</sub>* | 0.99 | 0.0 | 0.85 | 0.0 |

### B. Base Experiment i

The base experiment that employed five perfectly-measured populations over 201 days^10^ yielded an excellent solution in terms of model evolution and parameter estimates. Prior to examining the solution, we first plot the cost function versus the annealing parameter *β*, as this distribution can serve as a tool for assessing the significance of a solution.

Figure 3 shows the evolution of the cost throughout annealing, for the ten distinct independent paths that were initiated; the x-axis shows the value of Annealing

**Figure 3:**
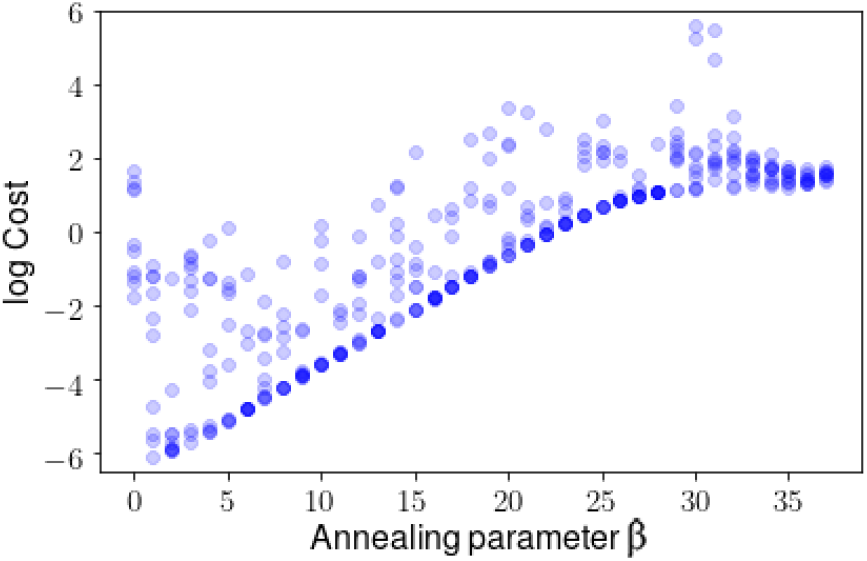
Cost function plotted at each annealing step *β* for the base experiment *i*, for ten paths in state space, where *β* scales the rigidity of the imposed model constraint. At low *β* the procedure endeavours to fit the measured variables to the simulated measurements. As *β* increases, the cost increases until it approaches a plateau (around *β =* 25), indicating that a solution has been found that is consistent with both measurements and model.

Parameter *β*, or: the increasing rigidity of the model constraint. At the start of iterations, the cost function is mainly fitting the measurements to data, and its value begins to climb as the model penalty is gradually imposed. If the procedure finds a solution that is consistent not only with the measurements, but also with the model, then the cost will plateau. In Figure 4, we see this happen, beginning around *β =* 25. Once *β* has reached 30, the plateau is established, with some scatter across paths. Unless noted, the reported estimates in this Section are taken at a value of *β* of 37: on the plateau. The significance of this plateau may become clearer once we examine the contrasting case of Experiment *ii*.

**Figure 4:**
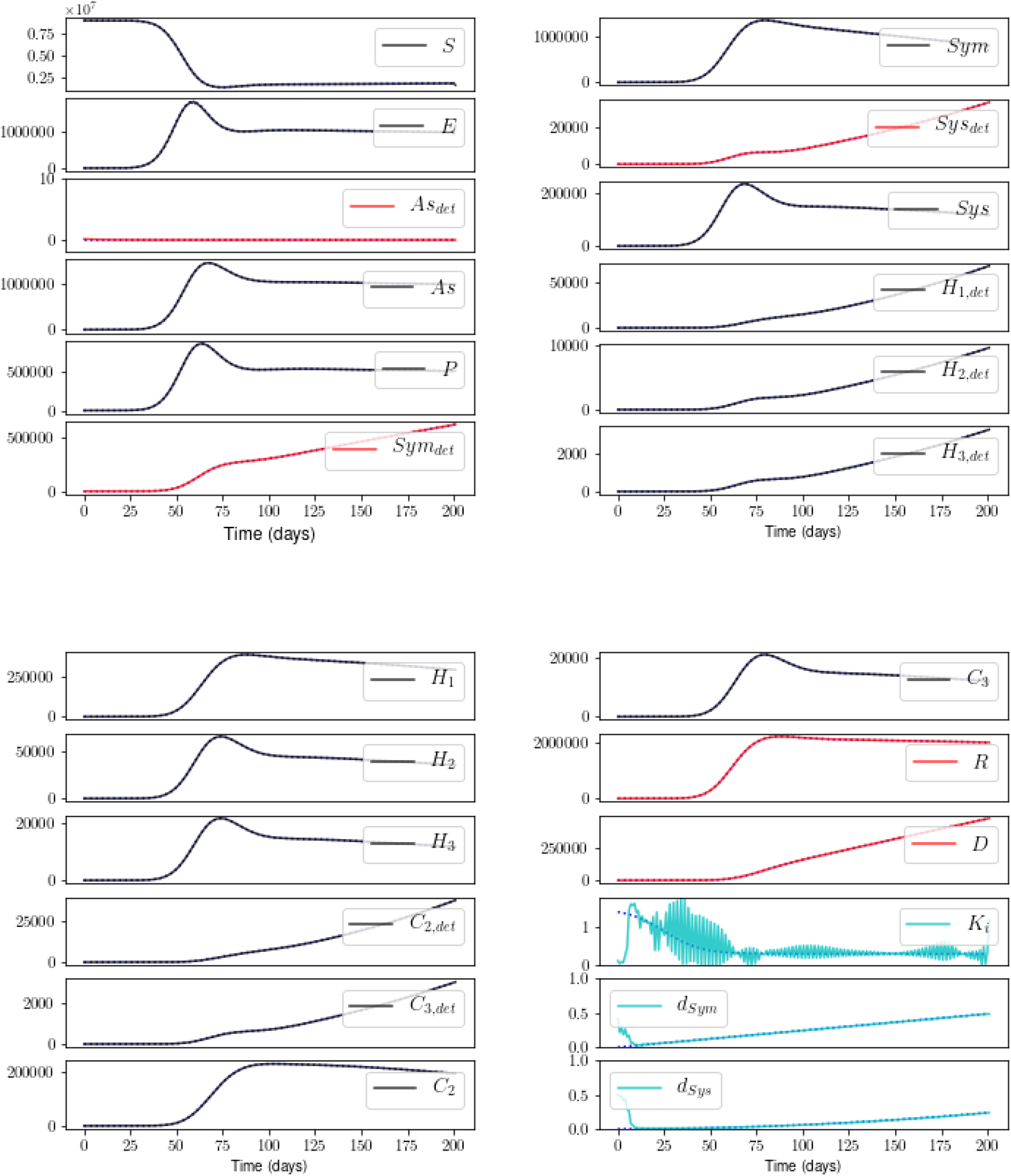
Estimates of the state - measured and unmeasured - variables, and the time-varying parameters *K_i_*, *d_Sym_*, and *d_Sys_*, for the base experiment *i*. Blue dotted: true model; black: estimate of unmeasured state; red: estimate of measured state; cyan: estimate of parameter. Excellent estimates of all states and parameters, except early values of *K_i_*; see text. Unless noted, all results reported in this Section are taken at a value for annealing parameter *β* of 37.

We now examine the state and parameter estimates for the base experiment *i*^11^. Figure 4 shows an excellent result, excepting *K_i_*(*t*) for times prior to its steep decline. We note no improvement in this estimate for *K_i_*(*t*), following a tenfold increase in the temporal resolution of measurements (not shown). The procedure does appear to recognize that a fast transition in the value of *K_i_* occurred at early times, and that that value was previously higher. It will be important to investigate the reason for this failure in the estimation of *K_i_* at early times, to rule out numerical issues involved with the quickly-changing derivative.^12^

### C. Experiment ii: no measurement of R

Figure 5 shows the cost as a function of annealing for the case with no measurement of Recovered Population *R*. Without examining the estimates, we know from the Cost(*β*) plot that no solution has been found that is consistent with both measurements and model: no plateau is reached. Rather, as the model constraint strengthens, the cost increases exponentially.

**Figure 5:**
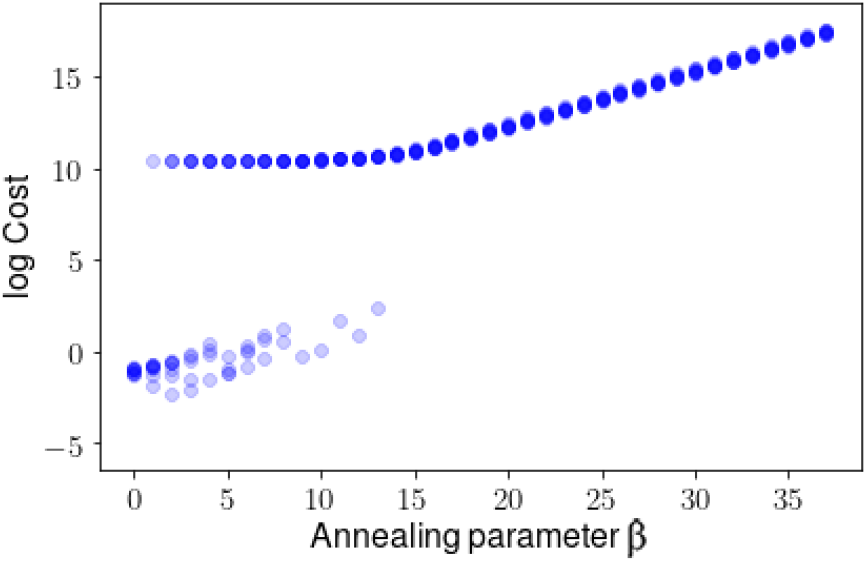
Cost versus *β* for Experiment *ii: R* is not measured. As *β* increases, the cost increases indefinitely, indicating that no solution has been found that is consistent with both measurements and model dynamics.

Indeed, Figure 6 shows the estimation, taken at *β =* 8, prior to the runaway behavior. Note the excellent fit to the measured states and simultaneous poor fit to the unmeasured states. As no stable solution is found at high *β*, we conclude that there exists insufficient information in *As_det_*, *Sym_det_, Sys_det_*, and *D* alone to corral the procedure into a region of state-and-parameter space in which a model solution is possible.

**Figure 6:**
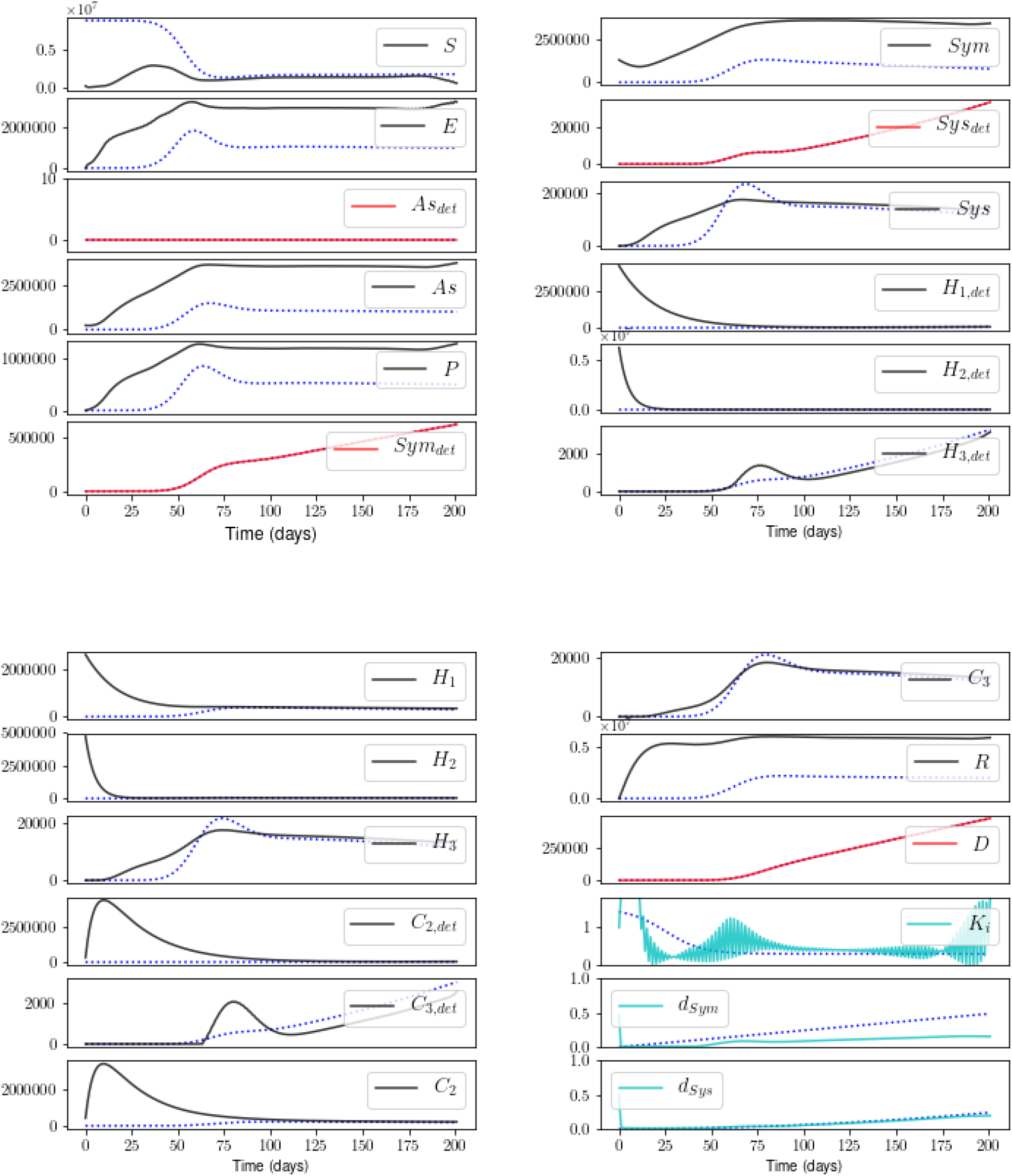
Estimates for Experiment *ii*. This result is taken at *β =* 8, prior to the exponential runaway in the cost. Estimates of unmeasured states and time-varying parameters are poor. For the key to the color-coding, see Caption of Figure 4.

### D. Experiment iii: shorter baseline of 112 days

Figure 7 shows estimates that include Measurement *R* but limit the temporal baseline to the first 112 days following time *t*_0_. The estimate suffers little from this reduction in the baseline of measurements. The only state estimate that is significantly different from the result shown in Figure 4 is *S* (the others are not shown), and the estimate of the transmission rate *K_i_*(*t*) is poor at all times. The estimates of the static parameters and trends in the detection probabilities are still captured well.

**Figure 7:**
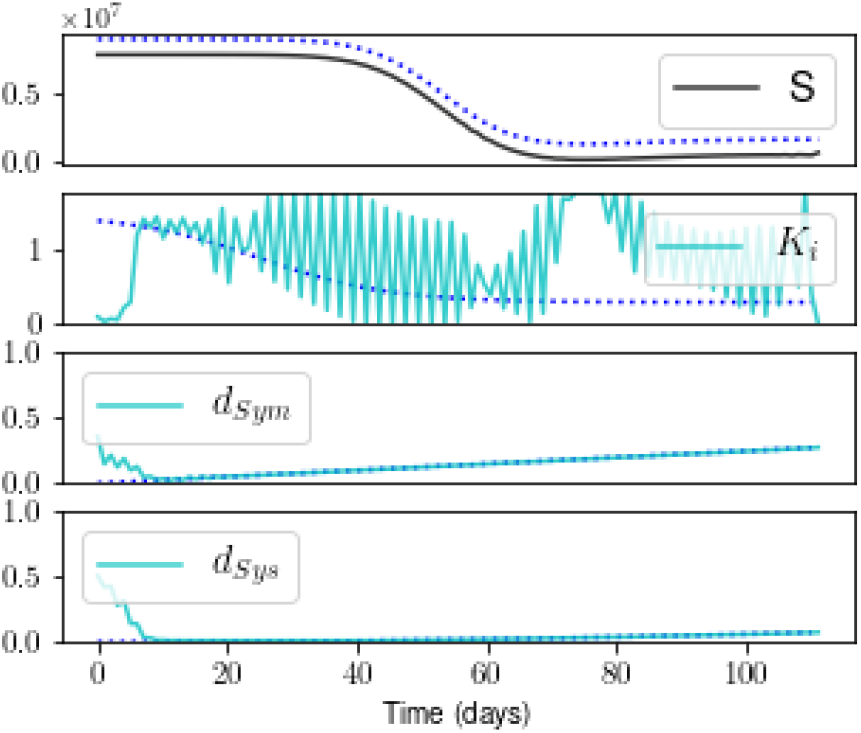
Estimates for Experiment *iii*, with a shorter baseline of 112 days. The state *S* is slightly underestimated, and the estimate of *K_i_* suffers at all times. The other states appear as they do in Figure 4 (not shown).

### E. Experiments iv and v: low noise added

With ~ 10% noise added to Measurement *R* (Experiment *iv*), a plateau appears on the cost-vs-*β* plot (Figure 8), indicating that a stable minimum has been found. Figure 9 shows that the noise propagates to the unmeasured States S, *E, As, P*, and to detection probability *d_sy_*_s_. The general evolution of the states and detection rates, however, is still captured well, with the one exception of *Sym*. Note that the low estimate for *Sym* is not offset by a high estimate for any of the other state variables. The addition of noise in these numbers, by definition, breaks the conservation of the population. In the future, we will ameliorate this problem by adding equality constraints into the cost function to impose the conservation of *N*.

**Figure 8:**
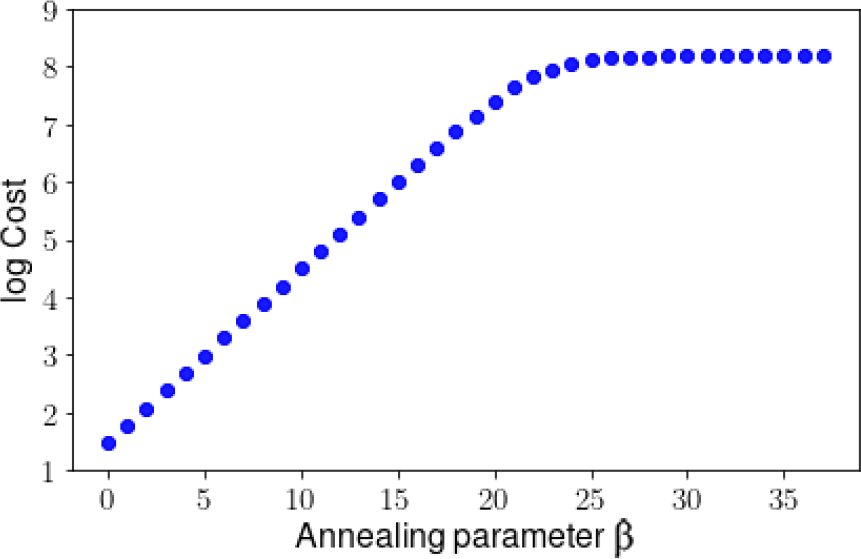
Cost versus *β* for Experiment *iv:* low noise added to *R*. A stable solution is found.

**Figure 9:**
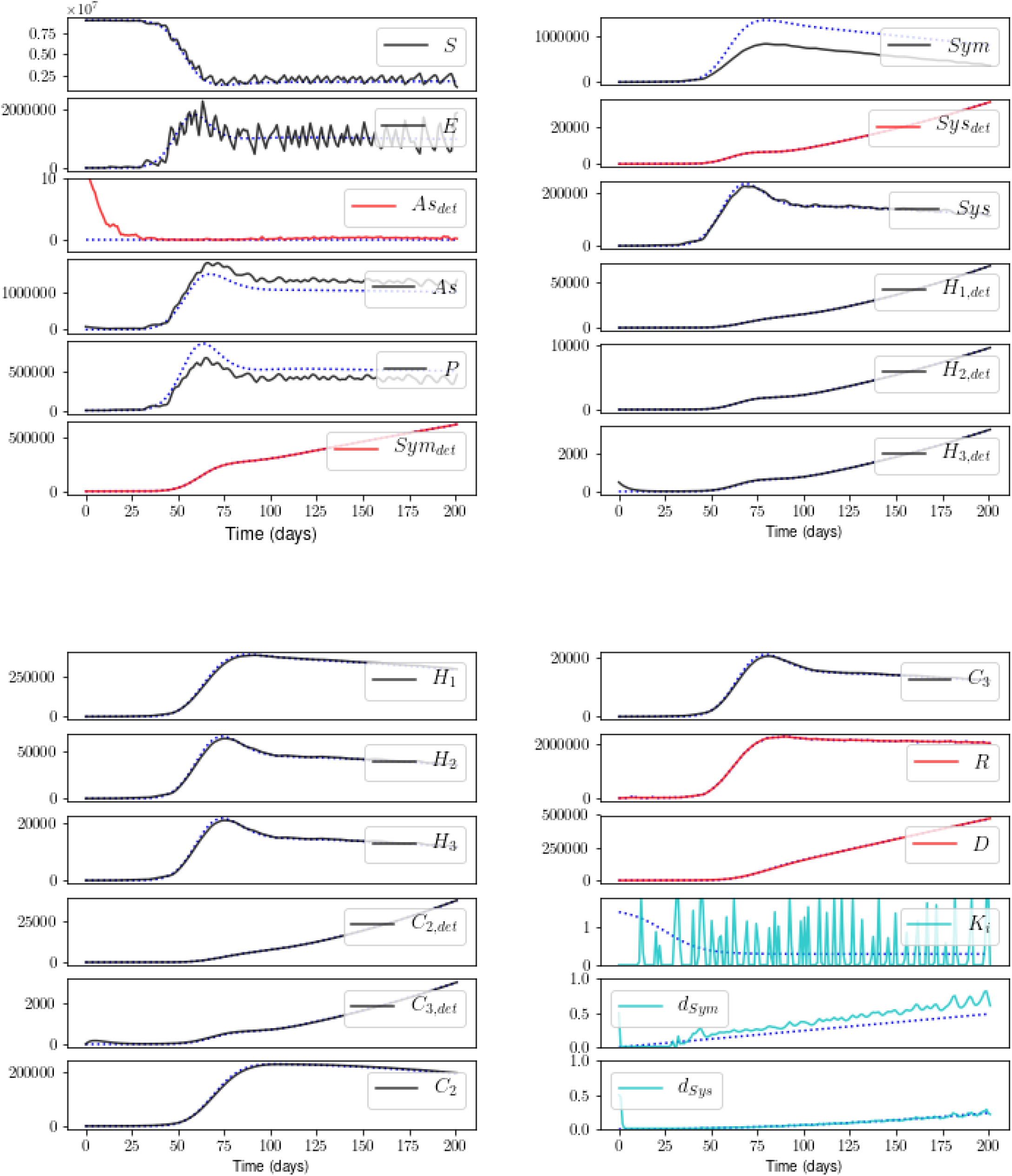
Estimates for Experiment *iv*. The noise added to *R* propagates to some unmeasured states and to the detection probability *d_Sym_*, but - with the exception of *Sym* - the overall evolution is captured well. The noise does preclude an estimate of transmission rate *K_i_*.

With the same noise level added to Measurements *Sym_det_* and *Sys_det_* as well (Experiment *v*), the state estimate is similar to that for Experiment *iv* (not shown). Estimates of the detection probabilities are poorer but still trace the general trends (Figure 10).

**Figure 10:**
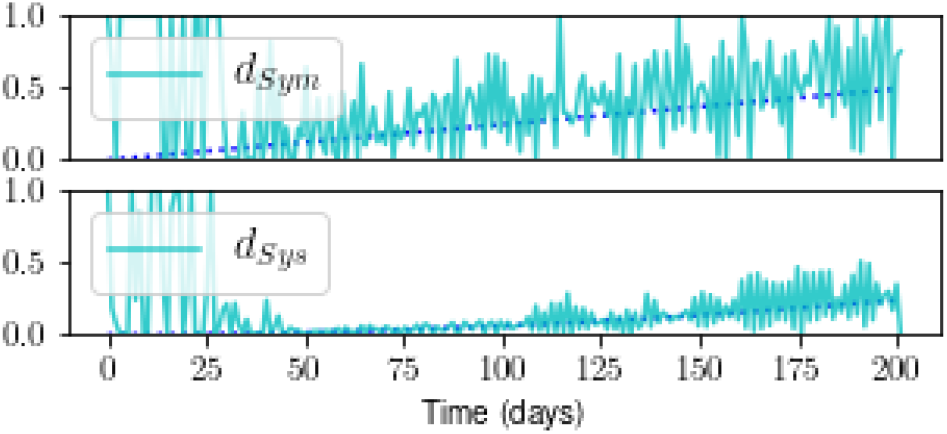
Estimates for Experiment *v*: noise added to *R*, *Sym_det_*, and *Sys_det_*. Estimates of detection probabilities are noisier than those for Experiment *iv*, although the trends are still captured. The state estimate is similar to that shown in Figure 9 for Experiment *iv* (not shown).

### F. Real data from Italy

We took the Confirmed (*C*), Recovered, and Dead cases in Italy that are listed in the github repository of the Johns Hopkins project Systems Science and Engineering [24]. These are cumulative numbers; *C* is the sum of R, D, and new cases. The *C* were divided among the model variables as described in *Experiments*.

Figures 11 and 12 show the result. The cost-vs-*β* plot indicates that a stable solution is found. The fits to the measured states are reasonable. The estimates of unmeasured states that impact hospital capacity (the *H* and *C* populations) appear smooth - and reasonable, with the exception of the features in *C*_3_*,_det_* and *H*_3_,*_det_* beginning at time *t*_0_. The estimates of *S* and *E*, on the other hand, are discontinuous. This may be due to the lack of a smooth solution for the time-varying parameters (not shown). We are currently amending the procedure to handle data that may include errors of unknown magnitude.

**Figure 11:**
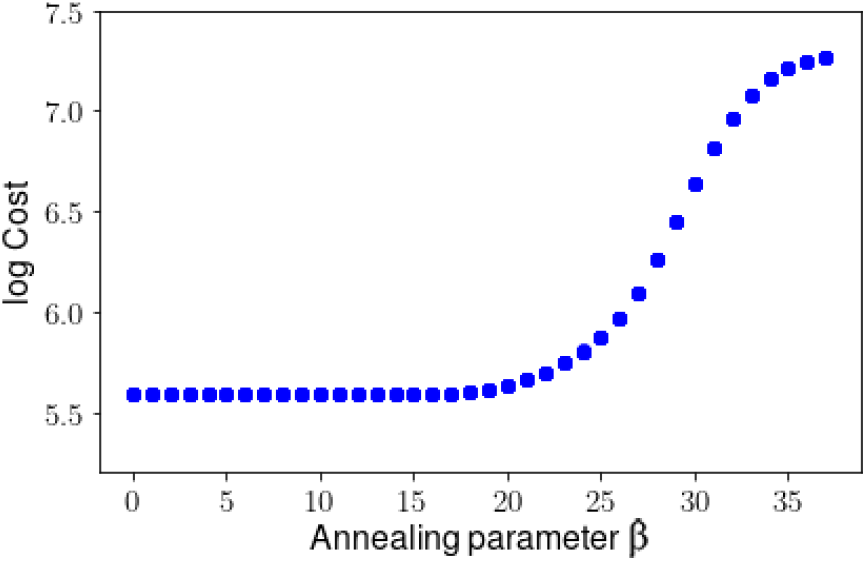
**Cost versus** *β* **using the Italy data**, indicating that a stable solution has been found.

**Figure 12:**
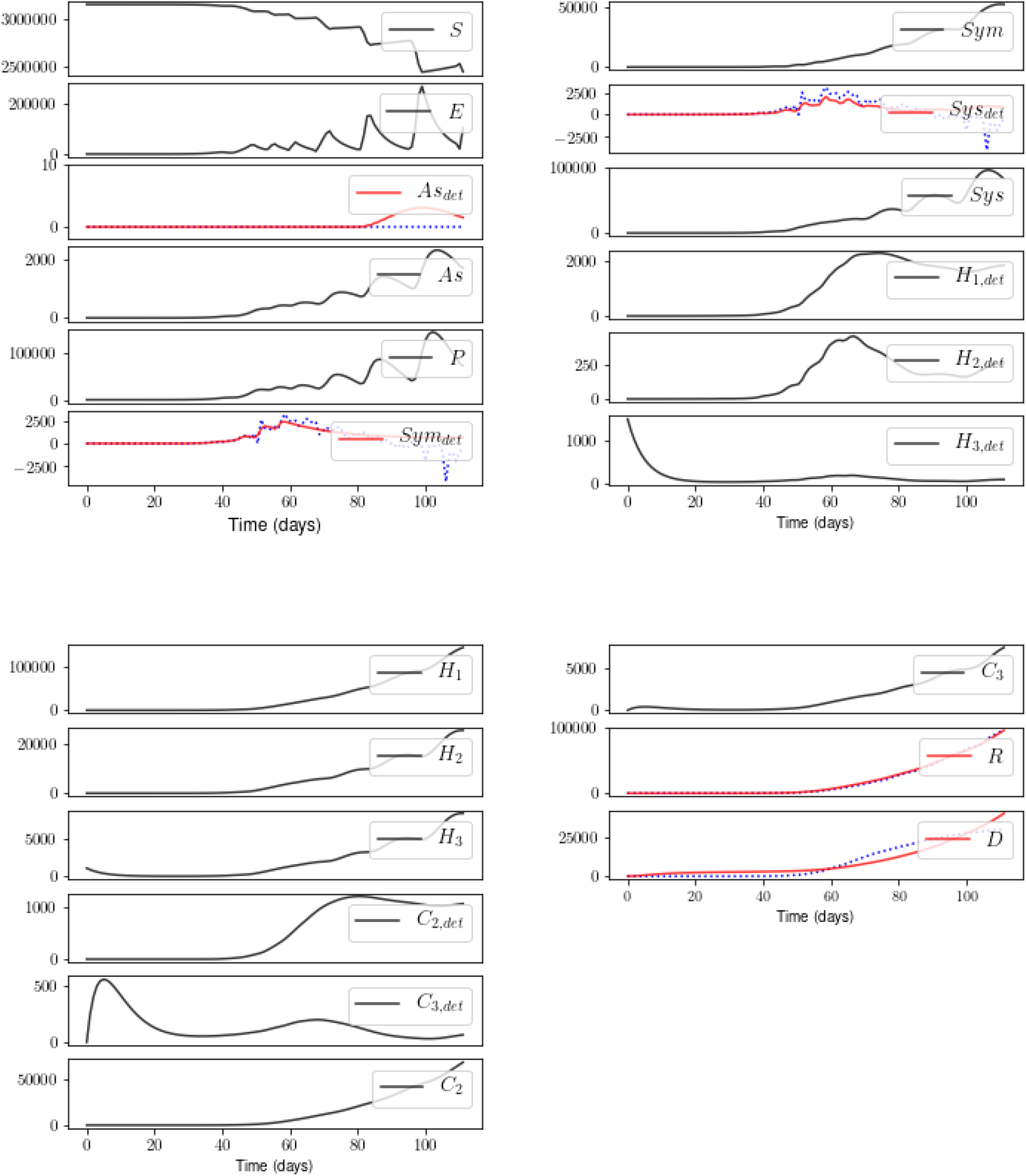
Estimates of all measured and unmeasured state variables, given the Italy measurements of *R*, *D*, and Confirmed *C* over 112 days. See text for comments and for our translation of *C* into state variables *As_det_, Sym_det_*, and *Sys_det_*.

## VI. CONCLUSION

We have endeavoured to illustrate how SDA can systematically identify the specific measurements, temporal baseline of measurements, and degree of measurement accuracy, required to estimate unknown model parameters in a high-dimensional model. We have used as our dynamical system an epidemiological model designed to examine the complex problems that COVID-19 presents to hospitals. In doing so, we have assumed knowledge of some model parameters. In light of these assumptions, we restrict our conclusions to general comments. We emphasize that estimation of the full model state requires measurements of the detected cases but not the undetected, provided that the recovered and dead are also measured. The model is tolerant to low noise in these measurements, excepting the transmission rate *K_i_*.

The SDA technique can be expanded in many directions. Examples include: 1) defining additional model parameters as unknowns to be estimated, including the fraction of patients hospitalized, the fraction who enter critical care, and the various timescales governing the reaction equations; 2) imposing various constraints regarding the unknown time-varying quantities, particularly transmission rate *K_i_*(t), and identifying which forms permit a solution consistent with measurements; 3) examining model sensitivity to the initial numbers within each population; 4) examining model sensitivity to the temporal frequency of data sampling.

The ultimate aim of SDA is to test the validity of model estimation, via prediction. In advance of that step, we are currently examining the stability of the SDA procedure over a range of choices for parameter values and initial numbers for the infected populations. As this pandemic is unlikely to abate soon, it is important to design well-informed plans for social and economic activity.

## Data Availability

All code and simulations will be made available via a github link. We generated no real data; only simulations.

## VII. ACKNOWLEDGEMENTS

Thank you to Patrick Clay for discussions on inferring exposure rates given social distancing protocols.

## Appendix A: Details of the model

**Table 3:**
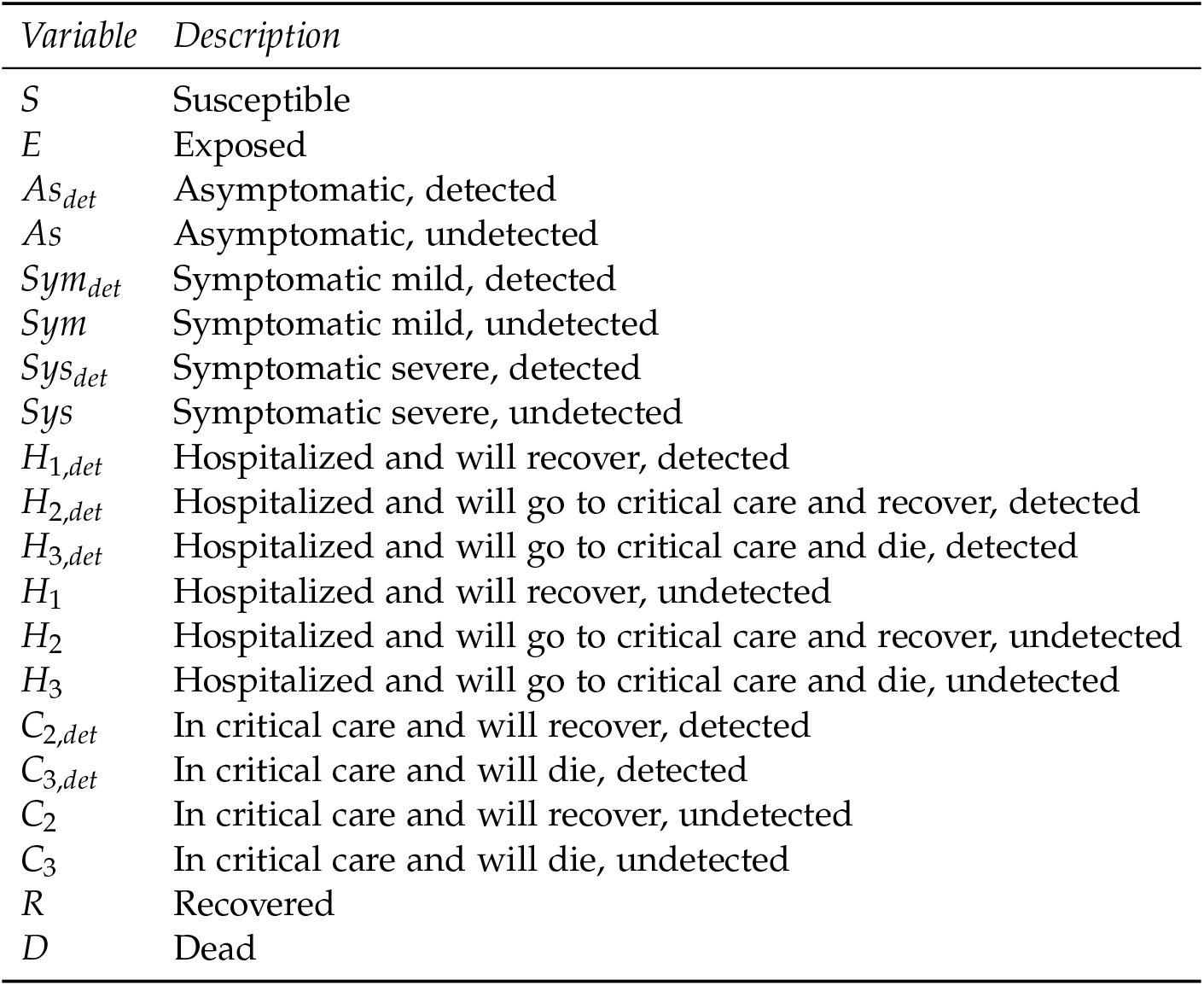
State variables of the COVID-19 transmission model. The “detected” qualifier signifies that the population has been tested and is positive for COVID-19.

### Reaction equations

The blue notation specified by overbrackets denotes the correspondence of specific terms to the reactions between the populations depicted in Figure 1. Note that the return of any portion of the *R* population to *S* is believed to be zero. Nevertheless, we assayed the ability of SDA to assimilate some portion of *R* back into *S*, as a proof-of-principle that it can be done readily.

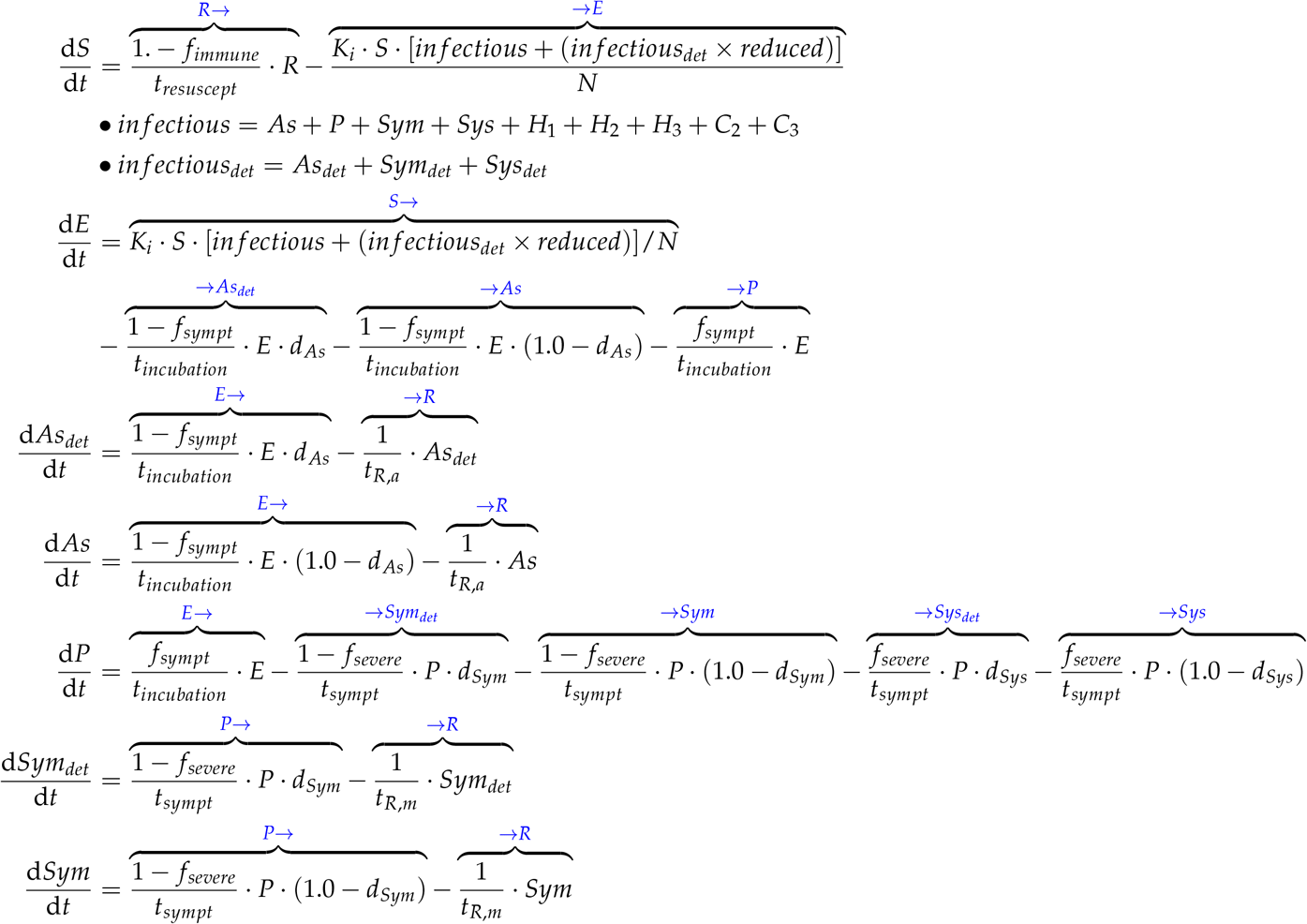

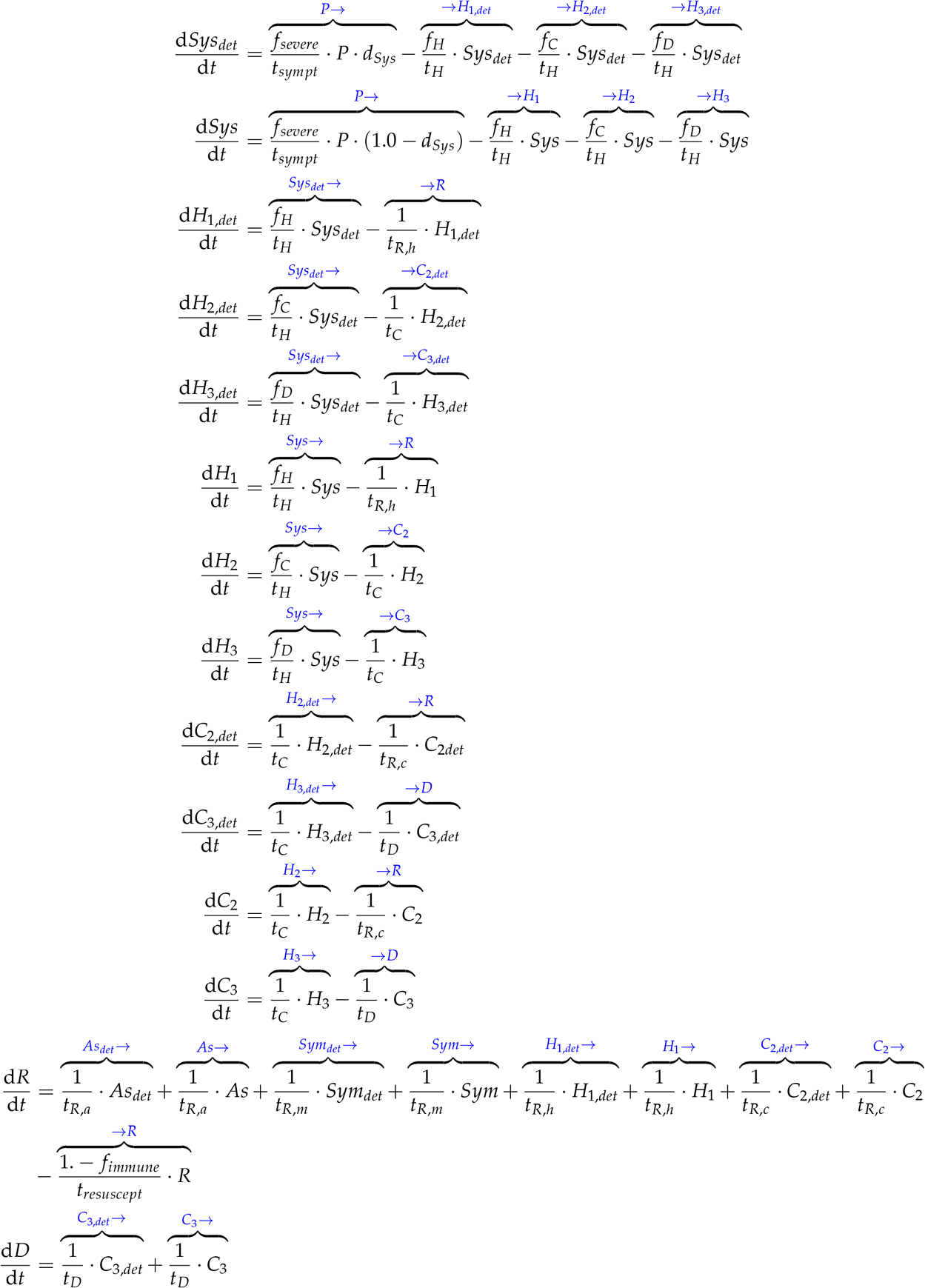

**Table 4:** The model parameters, with the unknown parameters to be estimated denoted in boldface. The unknown parameters *K_i_*, *_Sym_*, and *d_Sys_* are taken to be time-varying. The unknown parameters *f_sympt_* and *f_severe_* are taken to be intrinsic properties of the disease and therefore constant numbers. The detection probability of asymptomatic cases is taken to be known and zero. Units of time are days.

| Parameter | Description | Value |
| --- | --- | --- |
| $N$ | Total population | 9,000,000 |
| $f_{immune}$ | Fraction of recovered population that gains permanent immunity to re-infection | 0.9 |
| $t_{resuscept}$ | Time to re-enter the susceptible population, for those recovered who do not gain immunity | 1.0 |
| $reduced$ | The property that a detected case is likely to transmit less, via successful quarantine) | 0.5 |
| **$K_i(t)$** | **Transmission rate** | See *Appendix B* |
| $d_{As}(t)$ | Detection probability of asymptomatic cases | 0.0 |
| **$f_{sympt}$** | **Fraction of positive cases that produce symptoms** | **0.5** [40] |
| $t_{incubation}$ | Incubation time of disease | 5.0 |
| $t_{R,a}$ | Time to recovery for asymptomatics | 10.0 [42] |
| **$d_{sym}(t)$** | **Detection probability of mild symptomatics** | See *Appendix B* |
| **$d_{sys}(t)$** | **Detection probability of severe symptomatics** | See *Appendix B* |
| **$f_{severe}$** | **Fraction of symptomatics that are severe** | **0.3, likely an over-estimate** [41] |
| $t_{sympt}$ | Time to symptoms, for symptomatics | 5.0 |
| $t_{R,m}$ | Time from symptoms to recovery, for mild symptomatics | 20.0 [42] |
| $f_H$ | Fraction of severe cases that are hospitalized and then recover: $f_H = 1.0 - f_C - f_D$ | 0.6 |
| $f_C$ | Fraction of severe cases that require critical care and then recover | 0.3 [43] |
| $f_D$ | Fraction of severe cases that die | 0.1 |
| $t_H$ | Time from symptoms to hospital, for severe symptomatics | 5.0 |
| $t_{R,h}$ | Time from entering hospital to recovery, for severe symptomatics that do not require critical care | 20.0 [42] |
| $t_C$ | Time from entering hospital to critical care, for severe symptomatics | 5.0 [44] |
| $t_{R,c}$ | Time from entering critical care to recovery for severe symptomatics | 25.0 [42] |
| $t_D$ | Time from entering critical care to death, for severe symptomatics | 5.0 [45] |

## Appendix B: Unknown time-varying parameters to be estimated

The unknown parameters assumed to be time-varying are the transmission rate *K_i_*, and the detection probabilities *d_Sym_* and *d_Sys_* for mild and severe symptomatic cases, respectively.

The transmission rate in a given population for a given infectious disease is measured in effective contacts per unit time. This may be expressed as the total contact rate (the total number of contacts, effective or not, per unit time), multiplied by the risk of infection, given contact between an infectious and a susceptible individual. The total contact rate can be impacted by social behavior.

In this first employment of SDA upon a pandemic model of such high dimensionality, we chose to represent *K_i_* as a relatively constant value that undergoes one rapid transition corresponding to a single social distancing mandate. As noted in *Experiments*, social distancing rules were imposed in New York City roughly 25 days following the first reported case. We thus chose *K_i_* to transition between two relatively constant levels, roughly 25 days following time *t*_0_. Specifically, we wrote *K_i_*(*t*) as:

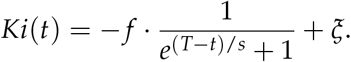

The parameter T was set to 25, beginning four days prior to the first report of a detection in NYC [35] to the imposition of a stay-home order in NYC on March 22 [39]. The parameter s governs the steepness of the transformation, and was set to 10. Parameters *f* and *ξ* were then adjusted to 1.2 and 1.5, to achieve a transition from about 1.4 to 0.3.

For detection probabilities *d_Sym_* and *d_Sy_*_s_, a linear and quadratic form, respectively, were chosen to preclude symmetries, and both were optimistically taken to increase with time:

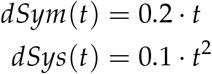

Finally, each time series was normalized to the range: [0:1], via division by their respective maximum values.

## Appendix C: Technical details of the inference experiments

The simulated data were generated by integrating the reaction equations (*Appendix A*) via a fourth-order adaptive Runge-Kutta method encoded in the Python package odeINT. A step size of one (day) was used to record the output. Except for the one instance noted in *Results* regarding Experiment *i*, we did not examine the sensitivity of estimations to the temporal sparsity of measurements. The initial conditions on the populations were: *S*_0_ *= N −* 1 (where *N* is the total population), *As*_0_ = 1, and zero for all others.

For the noise experiments, the noise added to the simulated *Sym_det_*, *Sys_det_*, and *R* data were generated by Python’s *numpy.random.normal* package, which defines a normal distribution of noise. For the “low-noise” experiments, we set the standard deviation to be the respective mean of each distribution, divided by 100. For the experiments using higher noise, we multiplied that original level by a factor of ten. For each noisy data set, the absolute value of the minimum was then added to each data point, so that the population did not drop below zero.

The optimization was performed via the open-source Interior-point Optimizer (Ipopt) [46]. Ipopt uses a Simpson’s rule method of finite differences to discretize the state space, a Newton’s method to search, and a barrier method to impose user-defined bounds that are placed upon the searches. We note that Ipopt’s search algorithm treats state variables as independent quantities, which is not the case for a model involving a closed population. This feature did not affect the results of this paper. Those interested in expanding the use of this tool, however, might keep in mind this feature. One might negate undesired effects by, for example, imposing equality constraints into the cost function that enforce the conservation of *N*.

Within the annealing procedure described in *Methods*, the parameter *α* was set to 2.0, and *β* ran from 0 to 38 in increments of 1. The inverse covariance matrix for measurement error (*R_m_*) was set to 1.0, and the initial value of the inverse covariance matrix for model error (*R_f_*_,0_) was set to 10^−7^.

For each of the five simulated experiments, ten paths were searched, beginning at randomly-generated initial conditions for parameters and state variables. For the Italy data, fifty paths were searched. All simulations were run on a 720-core, 1440-GB, 64-bit CPU cluster.

## Footnotes

1 Many details of this model, including compartmentalization by age and geographic region, are omitted from exploration in this paper.

2 It may interest the reader that one can derive this cost function by considering the classical physical Action on a path in a state space, where the path of lowest Action corresponds to the correct solution [26]

3 The model has been written to inform policy for the state of Illinois, but it generalizes to any geographical region.

4 For simplicity, we assume a closed population.

5 In the case of New York City, the first known symptomatic individual re-entered the U.S. from Iran several days prior, and may have been infected on that date [35].

6 The true number of initial cases is likely to be far higher. We are currently examining the model’s sensitivity to initial conditions on population numbers. Meanwhile, it is important to examine the SDA behavior over a range of choices for initial conditions, and we were struck by the rapid grown of the infected population - even given an initial infected population of one.

7 The reproduction number *R*_0_, in the simplest SIR form, can be written as the effective contact rate divided by the recovery rate. In practice, *R*_0_ is a challenge to infer [21, 36–38].

8 The full model permits *K*_i_ to vary as a function of multiple such rules, each initiated and rescinded at independent times.

9 The values of other model parameters also possess significant uncertainties given the reported data, including, for example, the fraction of those hospitalized that require ICU care. In future VA experiments, these quantities will also be treated as unknowns.

10 The implication of a 201-day requirement for accurate estimations would be a bleak outlook. We emphasize that these experiments are to be taken as *examples* for how the technique can be expanded to a host of realistic scenarios.

11 For all experiments, each solution shown is representative of the solution for all ten paths

12 As noted in *Experiments*, we chose *K_i_* to reflect a rapid adherence to social distancing at Day 25 following time *t*_0_, which then remains in place through to Day 201. For the form of *K_i_*, see *Appendix* B.)

